# Concordance between upper and lower airway microbiota in children with Cystic Fibrosis

**DOI:** 10.1101/2024.11.30.24318234

**Authors:** Sedreh Nassirnia, Valentin Scherz, Gilbert Greub, Giorgia Caruana, Patrick Taffé, Katia Jaton, Sebastien Papis, Klara M. Posfay-Barbe, Anne Mornand, Isabelle Rochat-Guignard, Claire Bertelli, Sandra A. Asner

## Abstract

**Background:** Sputum is the sample to monitor the lower respiratory tract microbiota in cystic fibrosis (CF), but young patients often cannot expectorate. We hypothesized that throat swabs could reflect lower airway colonization and assessed the concordance of bacterial community composition between paired sputum and throat swab samples from children with CF.

**Methods:** The prospective longitudinal multicenter MUCOVIB cohort included 379 samples from 61 CF children. Using V3-V4 16S rRNA amplicon metagenomics, we compared bacterial community diversity and composition between sputum and throat swabs in the full cohort and in 11 patients with paired samples from the same visit.

**Results:** Sputum and Throat swabs exhibited similar bacterial diversity, regardless of the exacerbation status, and presented a substantial agreement for detecting pathogens (Cohen’s Kappa: 0.6). Differences in bacterial abundance were observed (p=0.001), but not presence/absence (p=0.098). Community typing revealed three distinct community types, with 86% of paired samples falling into the same cluster, highlighting the homogeneity between sputum and throat swabs microbiota. Network analysis demonstrated slight, non-random similarities in microbial interactions between sample types (ARI = 0.08 and 0.10). The average distance between samples collected from the same visit was shorter (0.505, ± 0.056 95%CI), compared to sputum (0.695, ± 0.017) or throat swab (0.704, ± 0.045) from the same patient collected during different visits.

**Conclusions:** Throat swabs can provide representative information on lower respiratory microbiota. Clinicians should collect throat swabs rather than relying on sputum samples from previous visits to guide antibiotic prescriptions in CF children unable to expectorate.

## 1. Introduction

Cystic fibrosis (CF) is characterized by chronic pulmonary exacerbations, primarily driven by persistent bacterial infections, that lead to progressive lung disease [1–3]. The evolution of CF’s respiratory tract microbiome - from an initially diverse population to one increasingly dominated by pathogenic bacteria such as *Staphylococcus aureus*, *Haemophilus influenzae*, and *Pseudomonas aeruginosa* - highlights the interest in precise microbiota monitoring, to identify turning points in disease progression such as *P. aeruginosa* colonization [4,5].

The Climax-Attack Model (CAM) provides insights into the microbial dynamics within the CF lung [6,7]. The model differentiates ‘attack’ communities present during exacerbations, characterized by transient species colonization that triggers acute immune responses and lower pH through sugar fermentation, from ‘climax’ communities of more stable, slower-growing microbes resistant to antibiotics, associated with nitrogen waste management and an increased pH during stable periods. This framework underscores the dynamic nature of CF pathology and the importance of understanding microbial shifts for effective disease management [8]. Indeed, guidelines recommend regular microbiological workup to tailor antimicrobial therapy [10].

A significant challenge in CF management is obtaining the appropriate sample for respiratory microbiota analysis [9], especially among non-expectorating infants and young children. Bronchoalveolar lavage (BAL) is the gold standard and the best representative sample of lower respiratory microbiota. Yet, regular bronchoscopy is not recommended due to its invasive nature and the absence of proven clinical benefit [10]. Expectorated sputum stands as the best alternative for adults and older children capable of expectorating [10]. Throat swabs (TS) are the recommended alternative for infants and preschoolers [11]. Yet, culture-independent analyses have questioned the ability of TS to capture the lower respiratory tract microbiota, where chronic infection and inflammation occur [12]. Furthermore, interpreting TS results in children is complicated by age-related variations in the expected healthy microbial within different niches. This variability particularly affects infants and preschoolers, leading to inconsistent results across pediatric age groups and fueling debates about the reliability of TS in these cohorts [13–17].

The **Muco**viscidosis, Respiratory **V**iruses, and **I**ntracellular **B**acteria (MUCOVIB) project, a Swiss prospective longitudinal multicenter cohort, collected TS, sputum, nasopharyngeal swabs, bronchoalveolar lavage, and bronchial aspirate, to detect respiratory bacteria and viruses in 61 children with CF under 18 years of age [18, 19]. The efficacy of the 16S assay for pathogen detection compared to conventional culture results on respiratory samples showed fair agreement (0.4) based on Cohen’s Kappa score [23]. However, the 16S assay was more sensitive in detecting *H. influenzae* and *S. aureus* in respiratory samples compared to standard culture methods.

The present sub-study aims to clarify whether TS is a reliable proxy to sputum samples to reflect the microbial dynamics in the lower respiratory tract. By analyzing 16S rRNA sequence data from paired sputum and TS, we evaluated the concordance between microbial communities in these sample types, and key factors influencing their composition. Through community typing and ecological network analysis, focusing on keystone species and their interactions, we investigated the similarity in microbial community structure between sputum and TS during follow-up visits and pulmonary exacerbations.

## 2. Materials and methods

### 2.1. Study population and design

Details of the collection of clinical information, study design, and protocol have been published in the first MUCOVIB studies [18, 19]. Briefly, 61 patients were recruited from the respiratory clinics of Lausanne and Geneva University Hospital Centers and followed routinely by their treating pulmonologists. Culture, quantitative PCRs, and 16S amplicon-based metagenomics were used to assess the presence of viruses and bacteria in the respiratory tract. Overall, 269 TS and 51 sputa samples were collected longitudinally every 3 months from children with CF. The present MUCOVIB substudy evaluates the microbial composition of sputum and TS collected from the entire cohort and a subset of eleven individuals who provided 22 paired sputa and TS samples at the same visit.

### 2.2. Bioinformatics and statistical analyses

The composition of the microbial community was assessed through analyses of the V3-V4 regions of the 16S rRNA gene, sequenced using the Illumina MiSeq platform, as previously described [18, 19]. Raw sequencing reads are available in the European Nucleotide Archive (ENA) with project number PRJEB41059. The zAMP pipeline, an in-house DADA2-based bioinformatics tool, was used to process the paired-end sequences from the 16S amplicons (https://github.com/metagenlab/zAMP release v 0.9.15).

To assess the concordance between sputum and TS for detecting pathogens, Cohen’s Kappa statistic was computed using R software (v4.2.2). The vegan [20], mia [21], and ggplot2 packages were used for downstream statistical analysis and visualization. Wilcoxon rank-sum test was used to compare bacterial alpha diversity across two sample types, using paired tests for matched samples. Beta diversity was assessed by Bray-Curtis distance, considering the abundance of bacteria, and Jaccard distance, accounting for their presence/absence. Homogeneity of group dispersion was calculated using the betadisper test and bacterial composition between sample types was compared using the permutational multivariate ANOVA (PERMANOVA) test with 999 permutations. Adjusted p-values obtained from the BH method and values below 0.05 were considered statistically significant.

To examine the effect of host factors on variation of microbial composition, distance-based redundancy analysis (db-RDA) was performed with Bray-Curtis and Jaccard distances using capscale [20]. The species profiles from paired sputum and TS were used for community typing using the Dirichlet Multinomial Mixtures model (DMM) [22] and Laplace approximation, which was applied to identify the optimal number of DMMs. For both analyses, missing metadata for FEV1% (percentage of predicted Forced Expiratory Volume) and LCI (lung clearance index) were imputed using the median, as it is robust against outliers and skewed data.

The NetCoMi [23] package was used to construct and compare networks graphically and quantitatively. For this analysis, we applied sparse Correlations for Compositional data (SparCC) correlation coefficients [24], considering species that were present in at least 50% of each sample type and selecting associations with SparCC > 0.5. To identify topology and structural differences between networks of sputum and TS, a differential network was constructed where only differentially associated taxa are connected with only significant (P < 0.05) and strongly positive (SparCC > 0.5) correlations.

## 3. Results

### 3.1. Characteristics of the study population

The microbial composition of sample types was compared across the entire MUCOVIB cohort, consisting of 61 children (average age: 7.4; IQR [3.7; 12.2]). This cohort was followed for up to 756 days, with a median number of 7 visits (IQR [5;8]), resulting in the collection of a total of 379 samples. Furthermore, the sub-study focused on 22 paired sputum and TS samples collected during the same visit from 11 children, which represents 18% of the cohort. Among these 11 patients, 36% (4 children) contributed a single pair of samples, while 64% (7 children) contributed two or more pairs.

While the sex ratio was slightly imbalanced towards males (59%) in the entire cohort, in the subset of patients who provided both sample types on the same day of the visit, males and females accounted for 72.7% and 27.3%, respectively **(Table 1)**. FEV1% was lower and LCI was increased during exacerbations. Although FEV1% did not show a significant difference between the entire cohort and the paired cohort during follow-up visits, the difference was significant during pulmonary exacerbations. Additionally, LCI values were increased in the cohort of paired samples. Patients in the paired cohort tended to be older, with an average age of 12.7 years (IQR: 9.7- 14.6), and exhibited more severe disease symptoms, as correlated to the observed increase in LCI values.

**Table 1.** Patients’ characteristics in MUCOVIB and the subcohort.

| Clinical parameters | Complete cohort<br>61 patients | Paired samples subcohort<br>11 patients | Significant differences (p-value) |
| --- | --- | --- | --- |
| **Gender** |  |  |  |
| Female | 25 (41.0%) | 3 (27.3%) | 0.9 |
| Male | 36 (59.0%) | 8 (72.7%) | 0.7 |
| **Age at inclusion (Years)** | 7.4 [3.7, 12.2] | 12.7 [9.7, 14.6] | 0.02 |
| **Comorbidities** | 55 (90.2%) | 11 (100%) | 0.04 |
| Pancreatic insufficiency | 50 (82.0%) | 10 (90.9%) |  |
| Allergic bronchopulmonary aspergillosis | 4 (6.6%) | 2 (18.8%) |  |
| Diabetes | 6 (9.8%) | 2 (18.8%) |  |
| Liver disease | 5 (8.2%) | 2 (18.8%) |  |
| Others | 16 (26.2%) | 4 (36.6%) |  |
| **Follow-up (days)** | 553 [456, 640] | 470 [441, 585] | 0.09 |
| **Genotype** |  |  | 0.9 |
| Heterozygous for DeltaF508 | 24 (39.3%) | 5 (45.5%) |  |
| Homozygous for DeltaF508 | 28 (45.9%) | 5 (45.5%) |  |
| Other than DeltaF508 | 9 (14.8%) | 1 (9.1%) |  |
| **Pulmonary exacerbation (N of events)** | 32 | 5 | 0.9 |
| **FEV1%** |  |  |  |
| Follow-up visits (n=210) | 91.0 [81.0, 99.0] | 90.0 [73.0, 95.0] (n=5) | 0.4 |
| Pulmonary exacerbation (n = 14) | 82.0 [58.0, 97.5] | 71.0 [58.0, 85.75] (n = 8) | 0.001 |
| **Lung clearance index (LCI)** |  |  |  |
| Follow-up visits (n=194) | 9.3 [8.2, 11.1] | 10.75 [9.87, 13.50] | 0.1 |
| Pulmonary exacerbation (n = 9) | 9.7 [8.0, 12.8] | 10.8 [10.4, 13.67] | 0.006 |

FEV<sub>1</sub>: Forced expiratory volume in 1 second
LCI: The lung clearance index

### 3.2. Comparability of sputum and throat swabs for pathogen detection

To assess the reliability of TS, we measured pathogen detection rates in paired sputum and TS specimens using 16S assay. Compared to sputum, which was considered the gold standard, the 16S assay on TS showed an overall sensitivity of 78% and a specificity of 98%. **Table 2** details the Cohen’s Kappa scores, along with sensitivity and specificity values for the detection of five common CF pathogens. The Kappa score indicates a variable level of agreement in detection rates between paired sputum and TS samples depending on the bacterial species, ranging from substantial agreement for *Achromobacter* (0.69) to no agreement for *Stenotrophomonas maltophilia* (0), likely due to the small sample size. For *S. aureus*, throat samples yielded a sensitivity of 94%, a specificity of 25%, a positive predictive value (PPV) of 81%, and a negative predictive value (NPV) of 50%. In the case of *P. aeruginosa*, TS demonstrated sensitivities, specificities, PPVs, and NPVs of 50%, 83%, 40%, and 88%, respectively. The sensitivity rates varied significantly between the two sample types, with sputum samples yielding higher sensitivities.

**Table 2.**
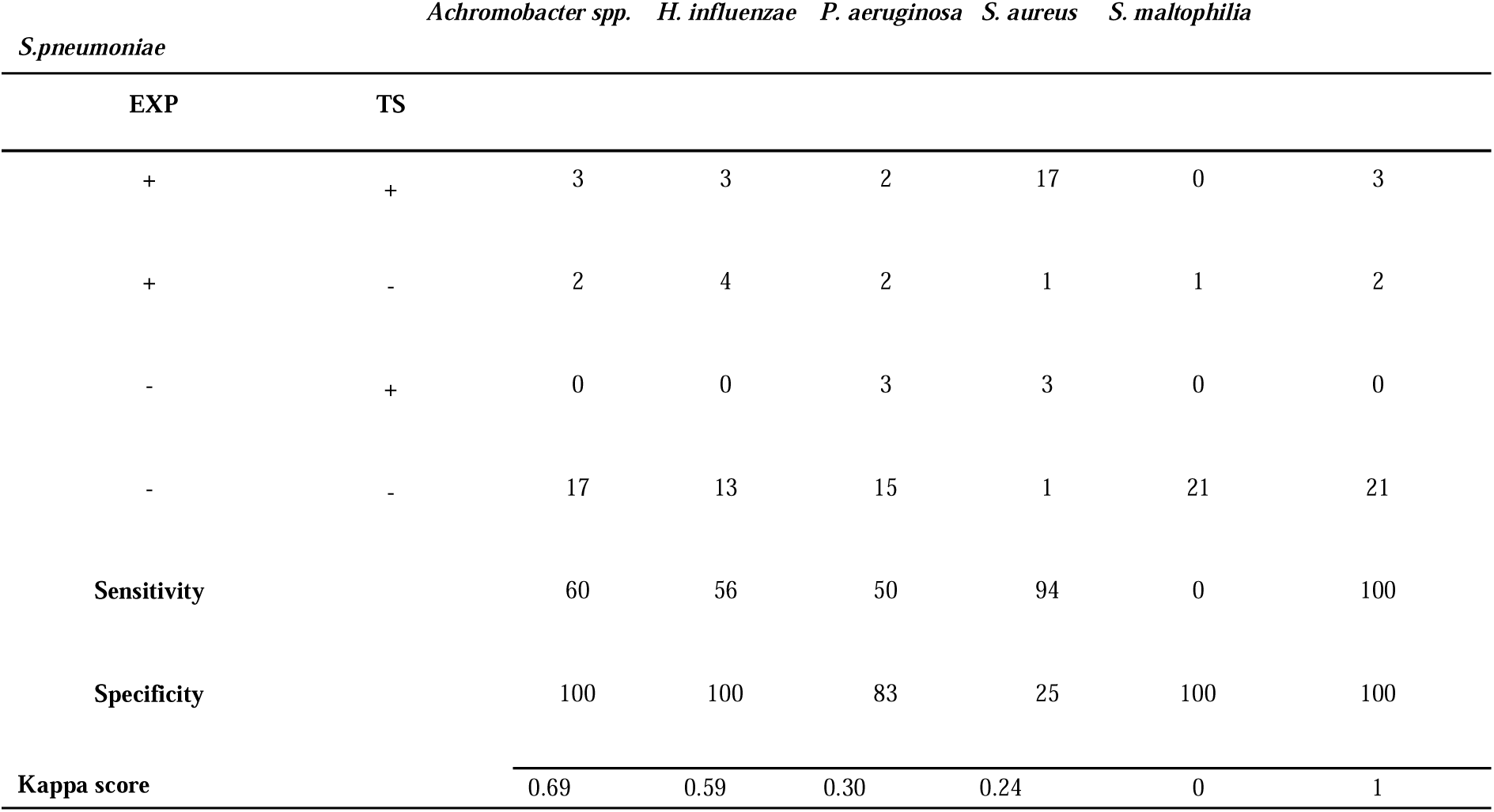
Sensitivity and specificity of throat swabs compared to sputum for pathogen detection by 16S amplicon-based metagenomics.

### 3.3. Microbiome diversity and composition in paired sputum and throat swab samples

Microbial diversity was calculated across sample types, using Chao1, indicating ASV richness, and the Shannon diversity index, which accounts for ASV richness and evenness **(Fig. 1A, B)**. Overall richness was not significantly different between the two sample types within the entire MUCOVIB cohort (p-value: 0.34) as well as in paired samples (p-values: 0.59). Similarly, comparisons of the Shannon index revealed no significant differences across both sample types. Hence, sputum and TS may offer similar insights into microbial diversity, irrespective of the patient’s exacerbation status **(Fig. S1A, B)**. Furthermore, bacterial communities across sample types were compared at the ASV level. In the MUCOVIB cohort, variability (dispersion) within microbial communities was significantly different between sputum and TS based on the abundance of bacteria (**Fig. 1C**, Bray-Curtis index, p-value: 0.05), but not based on the species composition (Jaccard index, p-value: 0.9). The significant difference in the overall bacterial community structure for both indices (PERMANOVA p-value: 0.001) indicated distinct microbial compositions between sample types. Similar results were observed within the paired samples, with a significant difference in dispersion using Bray-Curtis index (**Fig. 1D**, p-value: 0.04) but not with the Jaccard index (p-value: 0.41). In both cohorts, a PERMANOVA performed using sample type and visits (regular follow-up or exacerbations) as grouping factors, while stratifying by patient **(Fig. S1C, D)** suggested that both sample types (p-value: 0.001) and clinical status of the patient (p-value: 0.001) independently affect microbial community composition. However, their interaction did not show a significant impact (p-value: 0.64).

**Figure 1.**
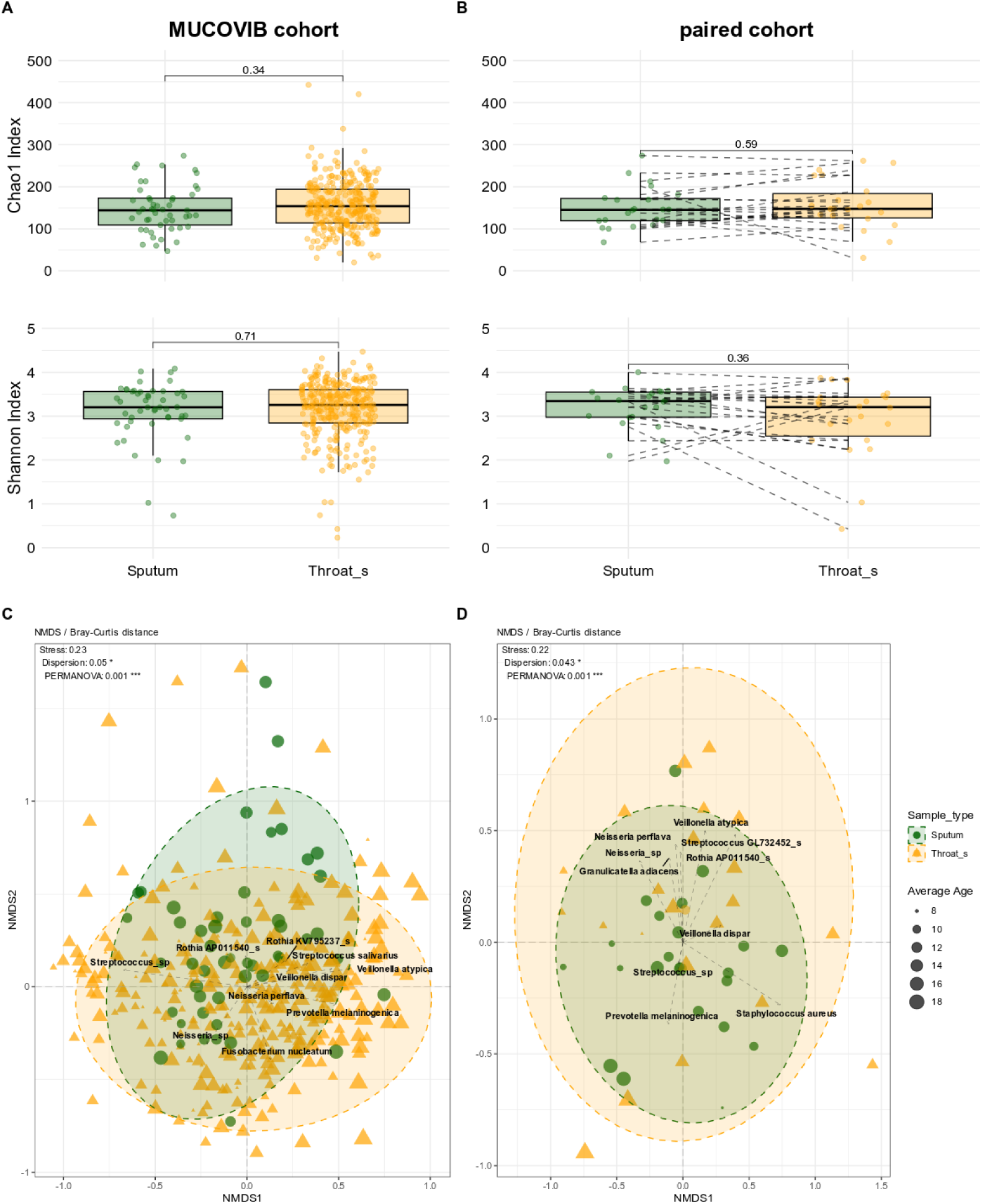
Microbiome diversity and composition in throat and sputum samples. **(A, B)** The box plot illustrates Chao1 and Shannon alpha diversity indices for each sample type in A. the MUCOVIB and B. paired sputum and throat swabs cohorts. The results of the Wilcoxon test (pairwise for paired samples) are indicated within the graphs. **(C, D)** Non-metric multidimensional scaling (NMDS) plot with Bray-Curtis distance at the ASV level for the complete and the sub-cohorts, respectively. Sputum samples were illustrated with green circles and throat swabs in orange triangles. Ellipses represent confidence intervals around clusters of sample types. Stress, dispersion, and PERMANOVA p-value indicate the goodness-of-fit, variability within groups, and the statistical significance of the observed clusters, respectively. Ten top abundant species are shown in the plots.

Patient-specific signatures in the microbiota were evident from the clustering of sputum and TS samples collected from the same visit observed in both the hierarchical clustering **(Fig. 2A)** and the NMDS (**Fig. 2B**) based on the presence/absence of the bacteria. A multivariate multiple regression model was built to evaluate the correlation between clinical and demographic covariates and the observed microbial communities. A significant proportion (57%, p-value: 0.001) of the variation in species abundances based on the adjusted R2 **(Fig. 2C)** was attributed to the patient’s visit. Only 15% of the total microbiota variation was explained by the exacerbation status, demographic factors (gender, age), and pulmonary functions (FEV1%, LCI) thereby suggesting that intra-individual variability mostly affected the microbial composition.

**Figure 2.**
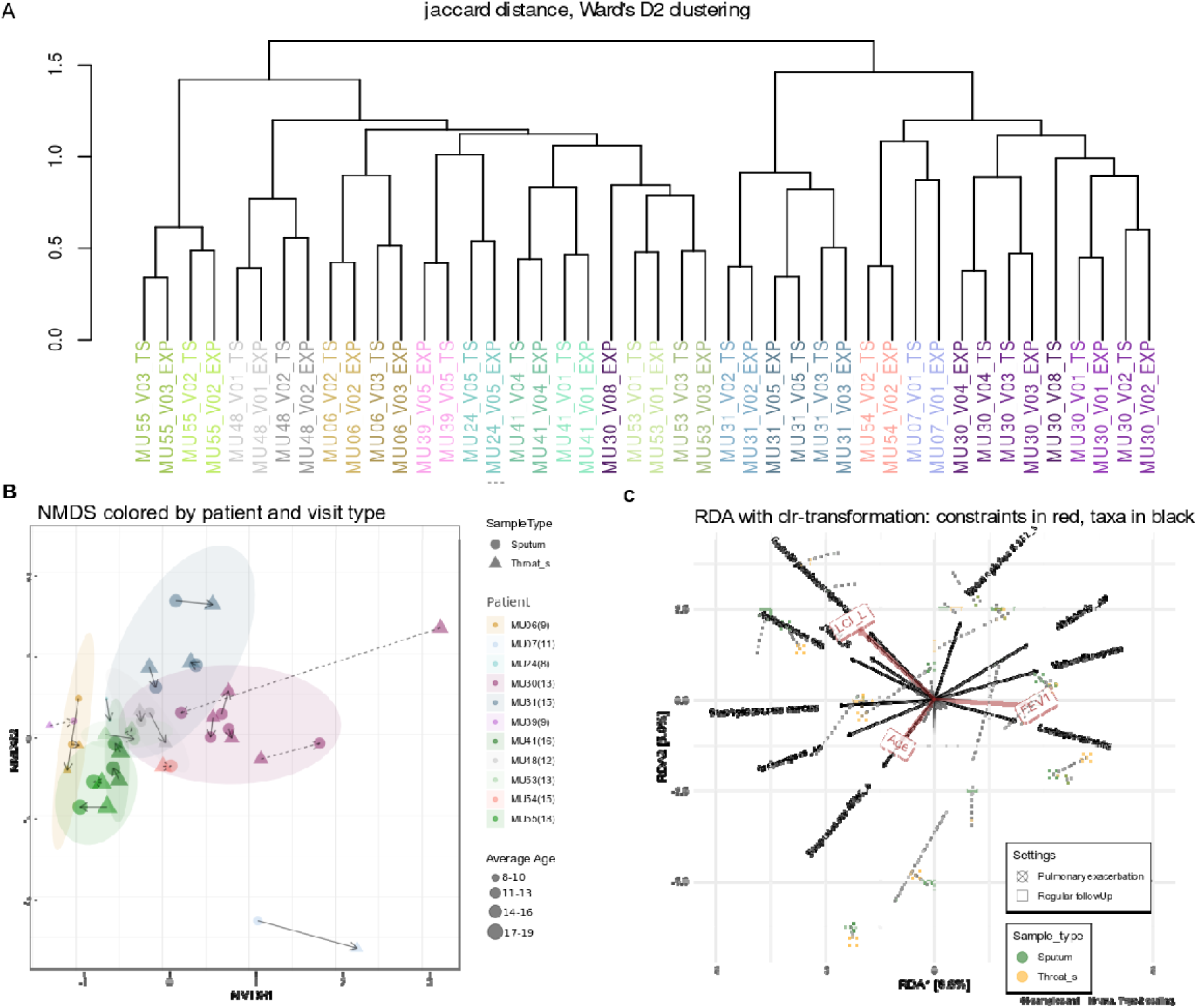
Patient-specific microbial community dynamics across visits. **(A)** The hierarchical clustering dendrogram based on Jaccard similarities illustrates the similarity in microbial community composition at the ASV level across different patient visits. Each leaf in the dendrogram represents a patient visit, with the color denoting individual patients and the shade variations reflecting different visits. The height of the branches indicates the degree of dissimilarity between clusters. **(B)** NMDS plot with Jaccard distance at the ASV level. Each point represents a sample colored by a patient and samples from the same visits connected by a line. The type of the line presents the kind of the visit, detailing individual variations in microbial communities from sputum and throat swabs during regular follow-up (solid lines) and pulmonary exacerbation (dashed lines) visits. Points are shaped according to sample types, with sizes representing the average age of patients. This visualization illustrates potential age-related shifts in microbial composition. To ensure anonymity, the age data was shifted by a random factor between 0 and 6 months. **(C)** Redundancy Analysis (RDA) using Bray-Curtis distance at the Species level. Each sample is represented with a shape indicating the type of the visit and its color represents the sample type. Dashed lines connect paired samples collected from the same visit. Arrows represent constrained variables with their direction and length indicating the strength and direction of their association with the microbial communities.

### 3.4. Microbial community typing

We then investigated whether variance in bacterial abundance reflects distinct microbial community types, or if community structures remain consistent despite quantitative variances across the two sample types. Using DMM, three distinct microbial community types, referred to as clusters, were detected **(Fig. 3A)**. We hypothesize that they could reflect host physiological states, rather than random statistical variances in microbial populations between sputum and TS samples, and potentially influence disease severity and clinical outcomes. Eighty-six percent of paired samples (19/22) were grouped into the same cluster, indicating a high degree of homogeneity between microbial communities in sputum and TS regardless of exacerbation status. The relative abundance heatmap of dominant taxa at species level provides a detailed view of the variability within each cluster **(Fig. 3A)**.

**Figure 3.**
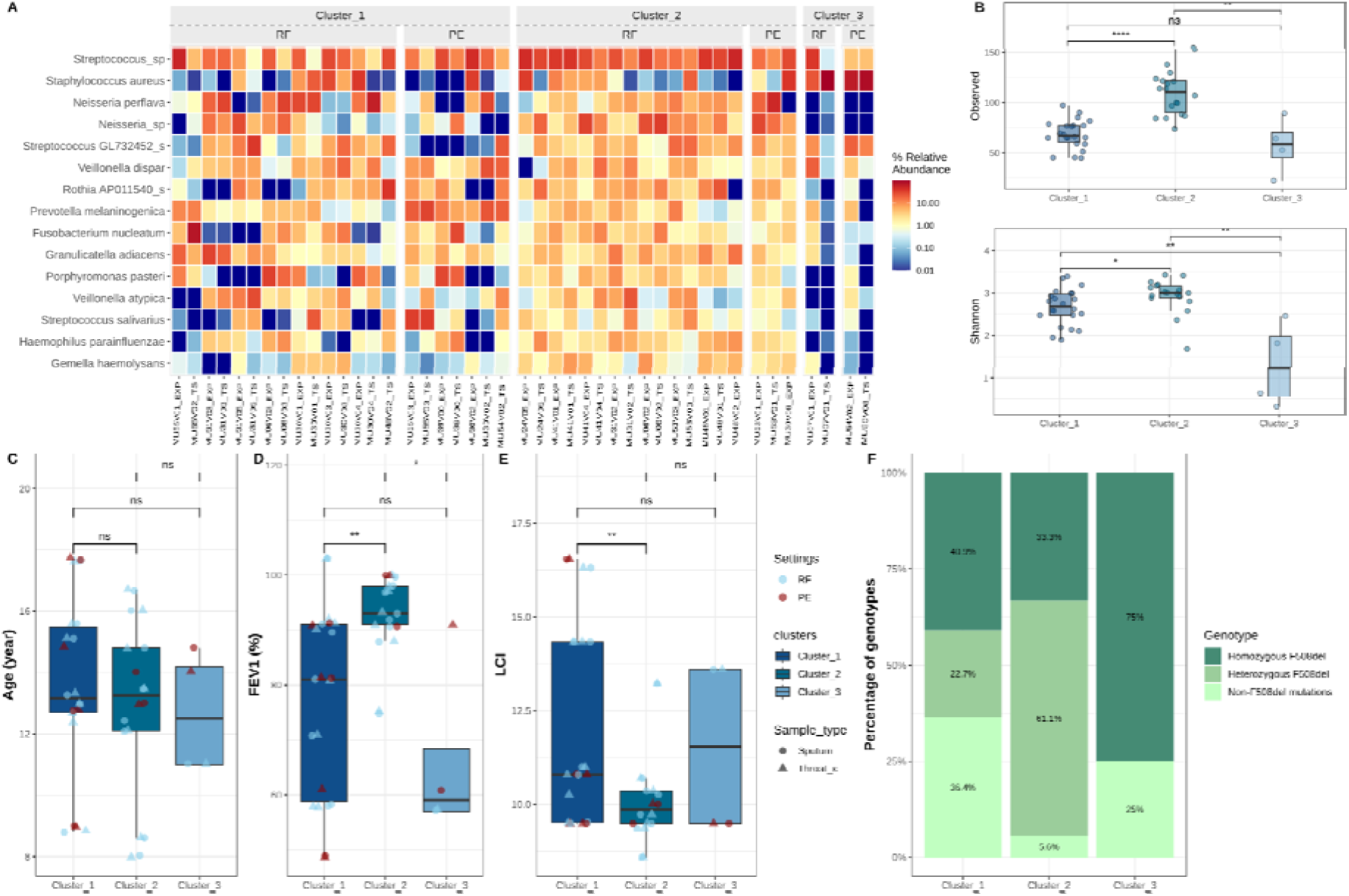
Community typing using Dirichlet multinomial mixture model (DMM) across sample types and clinical statuses. **(A)** Heatmap showing the relative abundance of the 15 most abundant species in the samples, grouped by clusters determined through DMM; Cluster 1 (n=10), Cluster 2 (n=8), Cluster 3 (n=4), with ‘n’ indicating the number of samples per cluster. Samples are further categorized by clinical status: Regular Follow-up (RF) and Pulmonary Exacerbation (PE). **(B)** Alpha diversity metrics, Observed species, and Shannon index, are compared across clusters. **(C)** Age distribution and **(D)** FEV1 percentage are presented for patients in each cluster. **(E)** Lung Clearance Index (LCI) values across clusters. **(F)** Distribution of CF genotypes within each cluster, showing proportions of Homozygous F508del, Heterozygous F508del, and Non-F508del mutations. Statistical significance is denoted as ns (not significant), * (p < 0.05), ** (p < 0.01), and **** (p < 0.0001), comparing the clinical statuses within each cluster.

Cluster 1 is notable for a community composition similar to the oropharyngeal flora, featuring a mix of facultatively anaerobic bacteria such as *Streptococcus* sp. and *Granulicatella adiacens,* alongside the obligate anaerobe *Fusobacterium nucleatum* and aerobic *Neisseria spp.*. This cluster presents a microbiota profile common to both throat and sputum samples, which may reflect a baseline microbial environment in CF patients. While sharing similarity with cluster 1 for the presence of anaerobic bacteria, *Streptococcus* sp. and *Neisseria* sp., cluster 2 is marked by a significant presence of *Veillonella* sp., a common organism from the oral microbiota, *Gemella haemolysans,* and *Haemophilus parainfluenzae*. Additionally, the abundance of bacteria present in cluster 2 is more stable over patient visits, whereas cluster 1 displays more dynamic changes. In contrast, cluster 3 is composed of samples from only three patients dominated by *S. aureus*.

Cluster 1, which contains more pulmonary exacerbation visits, displayed a lower species richness and evenness compared to cluster 2 **(Fig. 3B)**. Cluster 3 showed the lowest diversity, maybe owing to the small sample size (n=4) limiting the statistical power. When comparing patient clinical and demographic variables across clusters, there was no significant age difference **(Fig. 3C)**. Patients in cluster 1 had a lower FEV1% **(Fig. 3D)** and higher LCI scores **(Fig. 3E)** than cluster 2, indicating a reduced lung function. Clusters 1 and 3 had a higher proportion of patients with the homozygous F508del genotype, known to be associated with more severe disease phenotypes **(Fig. 3F)**.

### 3.5 Network

Expanding on the compositional similarities between sample types revealed by DMM analysis, we built an ecological network to delve deeper into the capacity of TS to capture microbial interactions within the lower respiratory tract microbiota of non-expectorating patients **(Figure 4A, B)**. Both networks displayed several clusters of microbial species that are associated, positively or negatively, within each sample type, exhibiting a mixture of conserved and niche-specific associations. For instance, the preservation of clusters of *Haemophilus*, *Veillonella*, *Porphyromonas*, *Neisseria*, and *Gemella* across both upper and lower respiratory samples in CF patients underlines the stability of these microbial associations despite the differing physical and immunological characteristics of these niches. Quantitative comparisons of global network properties between the two sample types revealed no significant difference in key metrics **(Table S1)**. Adjusted Rand Index (ARI) for the whole network and the Largest Connected Component (LCC) indicated a non-random similarity between the two networks (ARI of 0.08 and 0.10, respectively) with significant p-values (p=0.001 and 0.008). Whereas the sets of central nodes within the two microbial networks present moderate similarity in centrality measures, hub taxa presented a noticeable divergence (P(<=Jacc)=0.03*) between the two sample types **(Table S2)**. Furthermore, while several species are present in both networks, their roles as hub nodes differ, indicating the varied contributions of these central species to the ecological dynamics within each niche. In the sputum network, hub nodes such as *H. parainfluenzae*, *Solobacterium moorei,* and *Neisseria* sp. play central roles, while in the throat swab network, different taxa emerge as hubs, specifically *Capnocytophaga leadbetteri*, *Bergeyella* DQ241813_s, *Actinomyces* sp., and *Veillonella rogosae*. None of the nodes had significantly different centrality measures **(Table S1)** and no significant differential association was detected after multiple testing adjustments.

**Figure 4.**
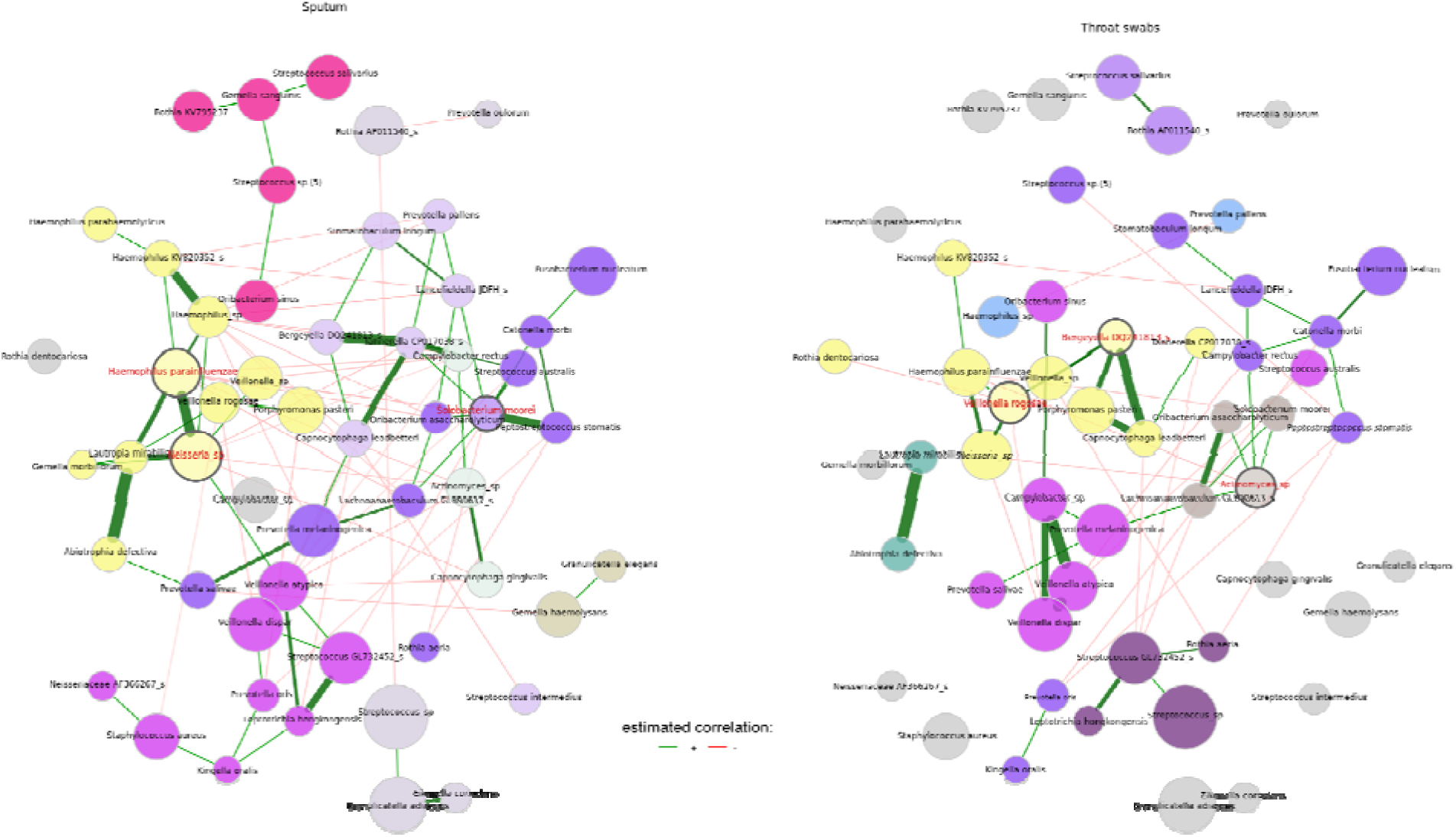
Microbial interactions network of sputum and throat swabs. The estimated correlations using SparCC are transformed into dissimilarities via the “signed” distance metric and the corresponding similarities are used as edge weights. The mclr-transformed abundance was used for defining hubs and scaling node sizes. Clusters were determined using greedy modularity optimization, and nodes were colored based on the clusters. To facilitate the comparison across sample types, the color of clusters differ between networks when the clusters do not share at least two nodes. Green edges correspond to positive estimated bacterial associations and red edges to negative ones. Only nodes that are unconnected in both networks were removed. To enhance visual comparison and highlight distinctions effectively, the layout computed for the sputum network was applied consistently across both networks. Hub species are highlighted with a red label.

To contextualize the ecological networks within the CAM framework and assess the ability of the TS to reflect exacerbation status, we constructed microbial networks at the climax and attack phases. In the climax phase, sputum samples exhibited a substantially larger relative LCC size (0.92) compared to TS (0.62), indicating a more complex and interconnected microbial network **(Table S3)**. Significant variation in hub taxa suggests that the species concentrating interactions in the lungs (represented by sputum) may not hold the same influence in the TS **(Table S4)**. During the attack phase, there was no substantial difference in the overall topology and the types of connections or graphlets between networks **(Table S5)**. However, a significant difference in the proportion of cooperative interactions among taxa (positive edge percentage) was observed between sputum (49.08%) and TS (54.13%). Similarly, network analysis identified significant differences according to exacerbation status **(Table S6**).

### 3.6. Temporal dynamics of microbial communities across patient visits

Building upon our analyses that demonstrate a concordance between microbial compositions of the upper and lower respiratory tracts in children with CF, we further investigated the temporal stability of these microbial communities. Specifically, we aimed to evaluate the reliability of TS as a proxy for lower respiratory tract microbiota at the time of patient visits, as compared to sputum samples or TS from previous visits. To address this, the Generalized Estimating Equations (GEE) method, which accounts for the intra-individual correlation of repeated measures, was employed to estimate the mean beta-diversity distances using both the Bray-Curtis and Jaccard indices while considering the dependence between observations from the same individual **(Fig. 5A, B)**. For the Bray-Curtis index, the GEE model indicated an average distance of 0.51 (95% CI: 0.45-0.57) between sputum and TS from the same visit. In contrast, the average distance between sputum samples from different visits was estimated at 0.57 (95% CI: 0.52-0.62), and the distance between TS samples from different visits was 0.66 (95% CI: 0.64-0.68), suggesting increased variability over time. Similarly, for the Jaccard index, sample types collected from the same visit presented a more similar bacterial composition (0.47; 95% CI: 0.42-0.52) than sputum or TS collected from different visits (0.68; 95% CI: 0.64-0.72 and 0.69; 95% CI: 0.67-0.70, respectively).

**Figure 5.**
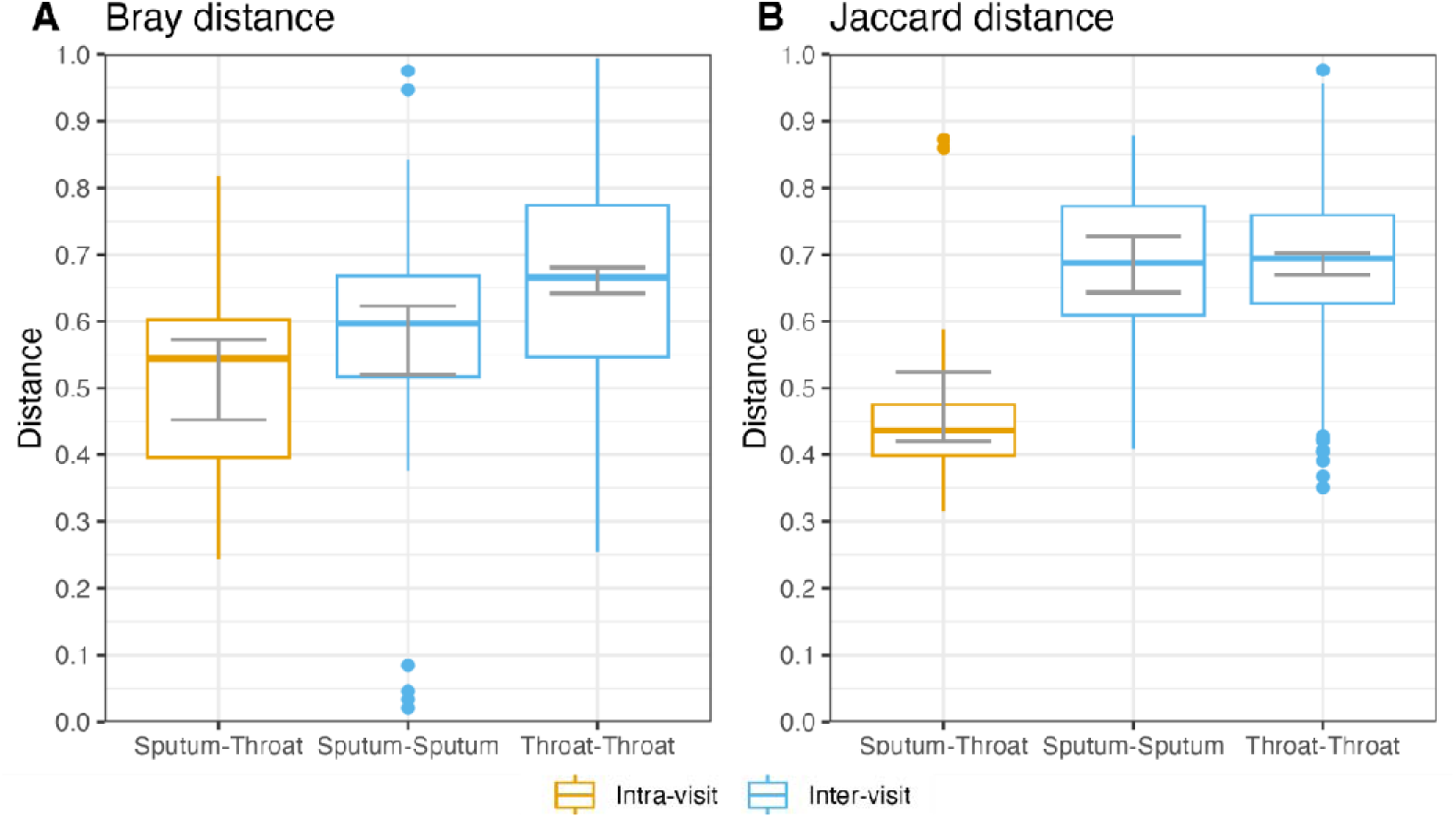
Beta-diversity distance between samples inter- and intra-visit. The average Bray-Curtis **(A)** and Jaccard **(B)** distances were calculated using GEE to account for intra-individual correlations. In both panels, orange boxes donate intra-visit comparisons, indicating the distance between throat swabs compared to sputum from the same visit, while blue boxes represent inter-visit comparisons, detailing the distance between samples from different visits. The median of each group is indicated by the central line within each box, with the box edges defining the interquartile range (IQR). The whiskers extend to the furthest points within 1.5 IQR from the box. Gray lines represent the 95% confidence intervals for each distance, providing insight into the variability and statistical certainty of the measurements.

## 4. Discussion

In this study, we compared the microbial landscape of TS and sputum using 16S rRNA amplicon sequencing to address a critical need for non-invasive, reliable sampling methods in CF. By focusing on paired samples collected on the same day and using a culture-independent method, we provided a direct comparison of the two microbiota and showed a reduced distance as compared to samples collected from the same site and the same patient during another visit. Our findings suggest the reliability of TS, collected on the same day of the visit, to mirror the CAM’s Climax and Attack phases in CF patients.

TS exhibited comparable microbial alpha-diversity (richness and evenness), with partially similar microbial composition to sputum. This is consistent with Boutin et al. [16], who demonstrated a close microbiota between sputum and TS in non-exacerbating patients. While both sample types harbor a broad range of microbial species, differences in relative abundances likely reflect anatomical and functional differences between the two niches.

The multivariate analysis showed that the predictive value of TS in relation to clinical and demographic covariates, compared to sputum, varies according to the individual-specific nature of the CF microbiota. Similarly, Prevaes et al. [12] noted that the degree of concordance between the microbial communities of different sample types with their corresponding BAL microbiota was variable within each patient when studied by either conventional culture or molecular assays.

Interestingly, community typing revealed that sputum-TS pairs often fall within the same cluster, suggesting consistency in microbial community structure across sample types. This consistency contrasts with the inter-individual variability observed across clusters, likely inherent to host-related factors such as CFTR F508del allele zygosity. Stephanie et al. [26] and Juan de Dios Caballero et al. [27] also observed unique microbiome patterns within each patient over time, despite variations in bacterial populations and without clear associations with exacerbation status or antibiotic treatment, thereby suggesting that personalized monitoring and treatment approaches may be beneficial in CF care.

Additionally, network analysis showed notable consistency in the microbial associations between sputum and TS across different clinical states (climax and attack phases) in our small population of children with exacerbation. The preservation of several anaerobes’ interactions, including *Veillonella* and *Prevotella*, between the two networks suggests they play roles in respiratory infections. Their ability to break down carbohydrates and mucins, could contribute to metabolic diversity, influencing immune modulation or biofilm formation within the mucosal environment of CF patients [28, 30, 31, 32]. The presence of these key bacterial taxa in both sample types support the potential utility of TS for monitoring microbial dynamics in young children with CF unable to expectorate and not yet colonized by *P. aeruginosa*. However, hub taxa—key players within each network—remained distinct, indicating differences in microbial roles across the two sample types.

Our 16S amplicon-based study provides insights into bacterial identity rather than function. Hence the taxa presence in TS may not necessarily reflect active biological roles specific to the ecological niche. Throat microbiota may partly be constituted of transient spillover bacteria from the mouth (saliva) and lungs (sputum) rather than a stable, throat-specific entity [29]. The larger Bray-Curtis distances between TS samples within individuals (**Fig. 6**) supported this hypothesis. Additionally, imbalanced sampling of paired samples among patients could introduce bias into the co-occurrence network. Patients with more frequent sampling may disproportionately influence inferred microbial associations and interactions. Not all multivariate analyses were adjusted to host-related factors such as F508del genotypes and antibiotic exposure, thereby limiting the assessment of specific patient-signatures.

Despite these limitations, the overall trends and key findings remain robust. When sputum samples are not available, the simplified collection of throat swabs has the potential to facilitate our understanding of disease progression and treatment response in young children with CF. Future studies with larger sample sizes including children with exacerbations should employ normalization techniques to weight samples or interactions based on the number of visits per patient. Large-scale longitudinal studies including F508del genotyping are needed to quantify the stability of microbial signatures, exchanges between compartments, and establish the functional profile of microbial communities across sample types.

## Data Availability

Raw 16S amplicon sequences are available in the European Nucleotide Archive (ENA) with project number PRJEB41059

