## Supplementary material for "Concordance between upper and lower airway microbiota in children with Cystic Fibrosis": Fig. S

**Supplementary figures and tables**

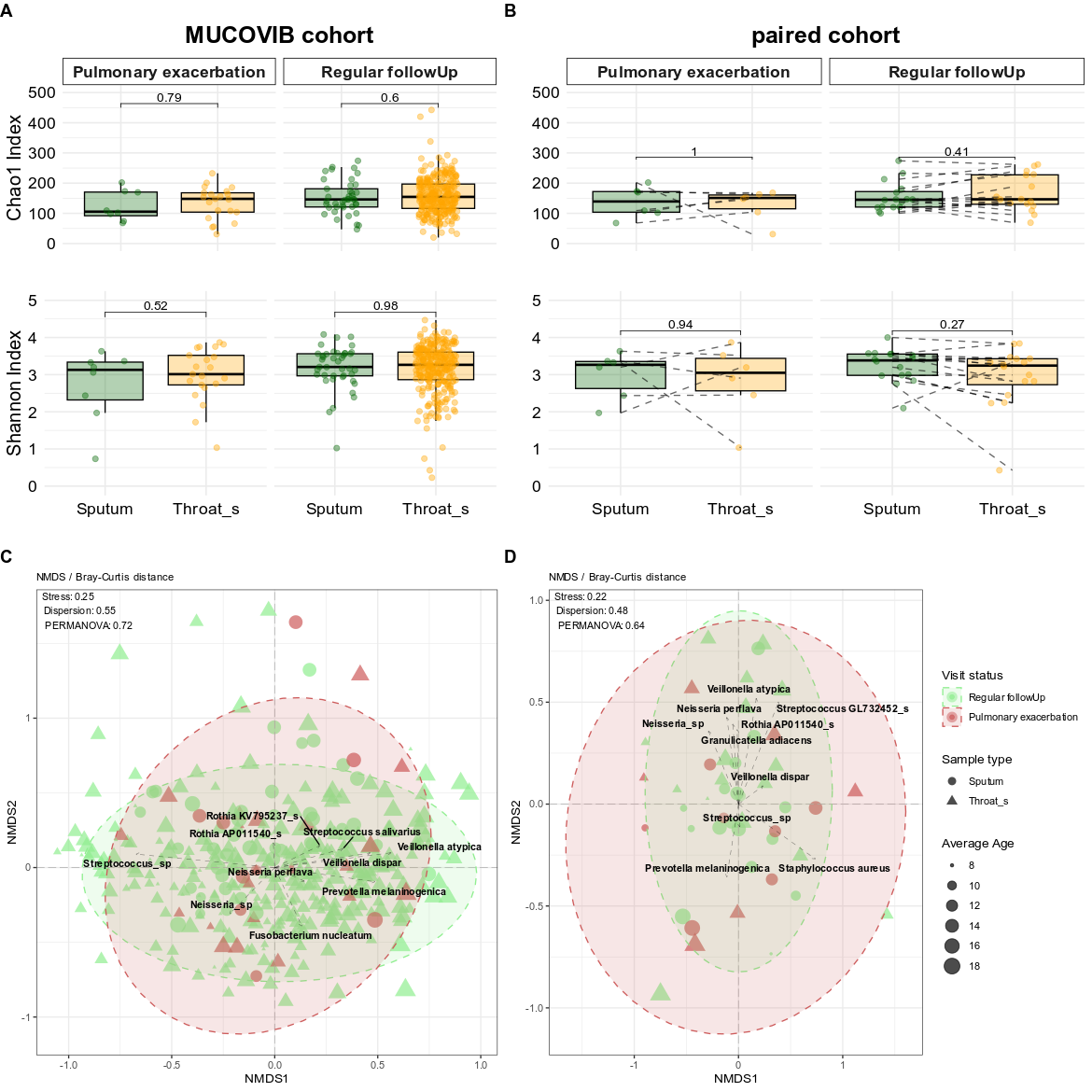

**Figure S1. Microbiome diversity and composition in throat and sputum samples during the regular follow up and pulmonary exacerbation.** The box plot illustrates Chao1 and Shannon alpha diversity indices for each sample type in **(A)** the MUCOVIB and **(B)** paired sputum and throat swabs cohorts. The results of the Wilcoxon test (pairwise for paired samples) are indicated within the graphs. Non-metric multidimensional scaling (NMDS) plot with Bray-Curtis distance at the ASV level for **(C)** the complete and **(D)** the sub cohorts, respectively. Sputum samples were illustrated with circles and throat swabs in triangles. Color code for samples and ellipses represent collected data during the regular visits in light green and pulmonary exacerbation in red. Stress, dispersion, and PERMANOVA p-values indicated in the plot. Ten top abundant species are shown in the plots.

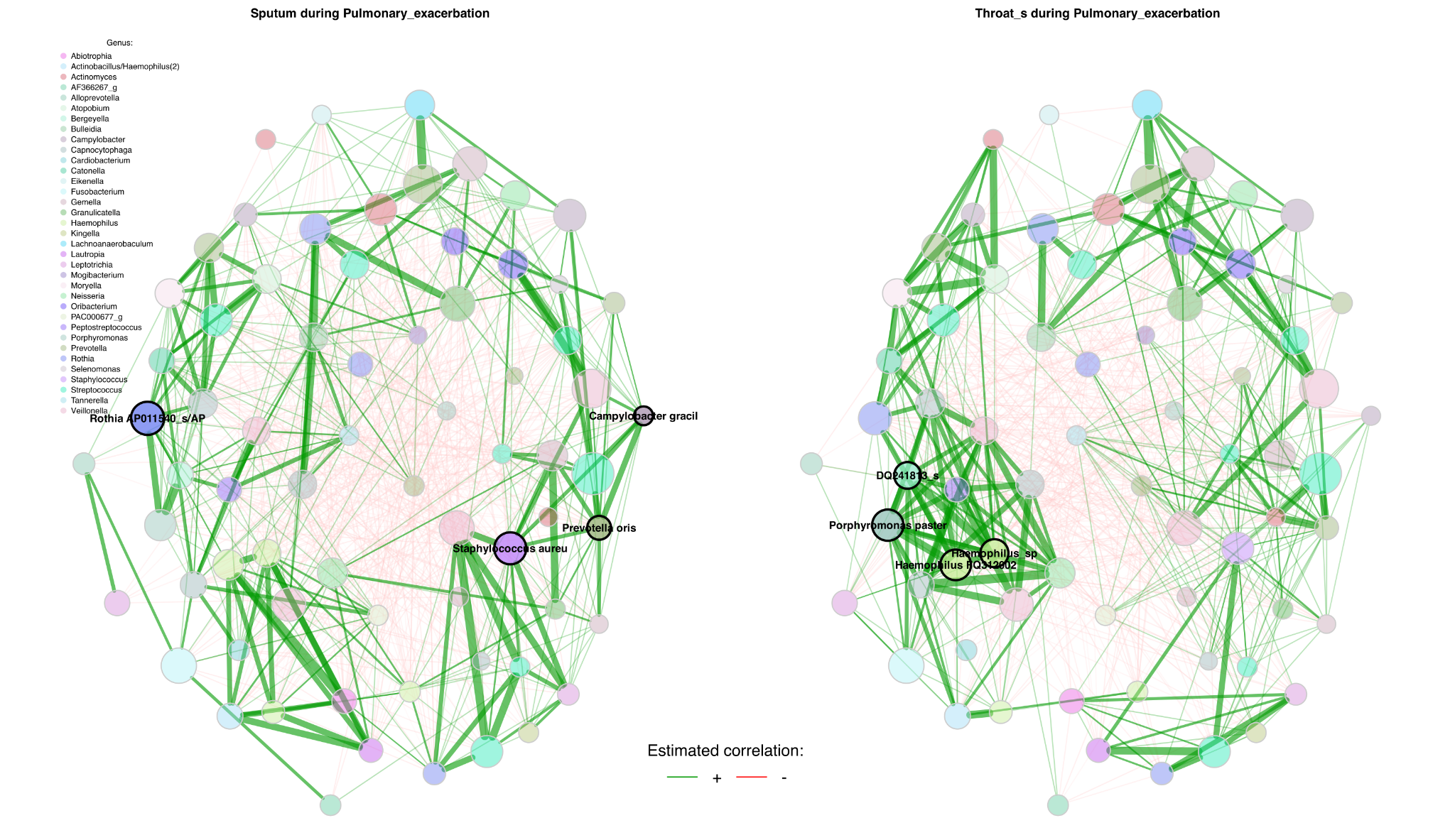

**Figure S2.** Microbial network of the attack phase

**Table S1. Sputum and throat swab networks quantitative comparison.** A comparative analysis of global network metrics and centrality measures across networks shown in Figure 4 was performed through permutation tests (1,000 permutations). The table highlights the top six species exhibiting the highest absolute differences in centrality measures measured for sputum and throat swabs. Centrality values are normalized to the range [0,1]. Centrality measures' P-values have been adjusted for multiple comparisons using the adaptive Benjamini-Hochberg method. Highly distinct eigenvector centralities, regardless of statistical significance, indicate bacteria that substantially differ in network plot between sputum and throat swab samples, such as *Neisseria* sp., which serves as a hub in sputum but is less central in throat swabs.

|  | **Sputum** | **Throat swab** | Abs diff. | Adj.p-value |
| --- | --- | --- | --- | --- |
| **Global network measures** | | | | |
| Relative LCC size | 0.70 | 0.58 | 0.12 | 0.28 |
| Clustering coefficient | 0.34 | 0.34 | 0.001 | 0.99 |
| Modularity | 0.33 | 0.48 | 0.14 | 0.34 |
| Positive edge percentage | 58.62 | 66.66 | 8.04 | 0.26 |
| Edge density | 0.10 | 0.09 | 0.01 | 0.67 |
| Natural connectivity | 0.03 | 0.04 | 0.006 | 0.04 |
| Vertex connectivity | 1.00 | 1.00 | 0.00 | 1.00 |
| Edge connectivity | 1.00 | 1.00 | 0.00 | 1.00 |
| Average dissimilarity* | 0.96 | 0.96 | 0.002 | 0.85 |
| Average path length | 1.79 | 2.04 | 0.24 | 0.64 |
| **Degree (weighted)** | | | | |
| *Neisseria* sp. | 3.69 | 1.37 | 2.31 | 0.83 |
| *Lautropia mirabilis* | 2.85 | 0.00 | 2.85 | 0.83 |
| *Staphylococcus aureus* | 1.61 | 0.00 | 1.61 | 0.91 |
| *Veillonella atypica* | 3.23 | 1.46 | 1.76 | 0.91 |
| *Haemophilus* sp. | 2.54 | 0.00 | 2.54 | 0.83 |
| *Veillonella rogosae* | 1.23 | 2.40 | 1.17 | 0.91 |
| **Betweenness centrality (normalized)** | | | | |
| *Solobacterium moorei* | 0.23 | 0.00 | 0.23 | 1 |
| *Lautropia mirabilis* | 0.17 | 0.00 | 0.17 | 1 |
| *Campylobacter rectus* | 0.45 | 0.20 | 0.19 | 1 |
| *Actinomyces* sp. | 0.01 | 0.17 | 0.15 | 1 |
| *Veillonella rogosae* | 0.08 | 0.22 | 0.13 | 1 |
| *Campylobacter* sp. | 0.00 | 0.12 | 0.12 | 1 |
| **Closeness centrality (normalized)** | | | | |
| *Haemophilus_sp* | 0.75 | 0.00 | 0.75 | 0.79 |
| *Lautropia mirabilis* | 0.74 | 0.00 | 0.74 | 0.61 |
| *Campylobacter_sp* | 0.00 | 0.76 | 0.76 | 0.16 |
| *Staphylococcus aureus* | 0.56 | 0.00 | 0.56 | 0.91 |
| *Fusobacterium nucleatum* | 0.00 | 0.69 | 0.69 | 0.79 |
| *Prevotella pallens* | 0.71 | 0.00 | 0.71 | 0.95 |
| **Eigenvector centrality (normalized)** | | | | |
| *Veillonella rogosae* | 0.16 | 1.00 | 0.83 | 0.99 |
| *Porphyromonas pasteri* | 0.03 | 0.82 | 0.78 | 0.99 |
| *Lautropia mirabilis* | 0.69 | 0.00 | 0.69 | 0.99 |
| *Haemophilus_sp* | 0.65 | 0.00 | 0.65 | 0.99 |
| *Solobacterium moorei* | 1.00 | 0.50 | 0.49 | 1.00 |
| *Campylobacter_sp* | 0.00 | 0.50 | 0.50 | 1.00 |

*: Dissimilarity = 1 - edge weight

**Table S2. Jaccard similarity for edge sets corresponding to the networks shown in Figure 4.** Jaccard index expresses the similarity of the sets of most central nodes and the sets of hub species between the two networks. This index ranges from 0 to 1, where 0 means no similarity and 1 means complete similarity. “Most central” nodes have an eigenvector centrality value above 95% quantile of the observed distribution of centrality scores. The p-values indicate the probability of obtaining a Jaccard index less than or equal to (<=Jacc) or greater than or equal to (>=Jacc) the observed value under the null hypothesis of no association.

| **Measure** | **jacc** | **P(<=Jacc)** | **P(>=Jacc)** |
| --- | --- | --- | --- |
| degree | 0.33 | 0.58 | 0.57 |
| betweenness centr. | 0.25 | 0.26 | 0.86 |
| closeness centr. | 0.33 | 0.58 | 0.57 |
| eigenvec. centr. | 0.38 | 0.77 | 0.35 |
| hub taxa | 0.00 * | 0.06 | 1.00 |

**Table S3. Network comparison in the climax phase**

|  | Sputum | Throat_s | Abs_diff. | P_value |
| --- | --- | --- | --- | --- |
| **Global network measures** | | | | |
| Relative LCC size | 0.90 | 0.62 | 0.27 | 0.004 ** |
| Clustering coefficient | 0.35 | 0.37 | 0.01 | 0.86 |
| Modularity | 0.31 | 0.40 | 0.09 | 0.37 |
| Positive edge percentage | 56.52 | 60.87 | 4.34 | 0.55 |
| Density | 0.02 | 0.03 | 0.01 | 0.12 |
| Vertex connectivity | 1.00 | 1.00 | 0.00 | 1.00 |
| Edge connectivity | 1.00 | 1.00 | 0.00 | 1.00 |
| Average path length | 2.07 | 1.83 | 0.24 | 0.28 |
| **Degree (weighted)** | | | | |
| *Tannerella* CP017038_s | 0.00 | 12.490 | 12.49 | 0.41 |
| *Haemophilus haemolyticus* | 9.67 | 2.103 | 7.57 | 0.98 |
| *Gemella haemolysan* | 8.57 | 2.680 | 5.89 | 0.98 |
| *Campylobacter gracilis* | 9.36 | 4.010 | 5.35 | 0.98 |
| *Haemophilus parainfluenzae* | 7.04 | 12.060 | 5.01 | 0.98 |
| *Eikenella corrodens* | 5.24 | 0.990 | 4.25 | 0.98 |
| **Eigenvector centrality (normalized)** | | | | |
| *Gemella haemolysans* | 0.78 | 0.05 | 0.72 | 0.65 |
| *Schaalia* JVLH_s | 0.00 | 0.61 | 0.61 | 0.65 |
| *Campylobacter gracilis* | 0.86 | 0.28 | 0.58 | 0.65 |
| *Haemophilus haemolyticus* | 0.64 | 0.10 | 0.54 | 0.65 |
| *Rothia* KV795237_s | 0.64 | 0.10 | 0.53 | 0.65 |
| *Streptococcus_sp* | 0.80 | 0.31 | 0.48 | 0.65 |

**Table S4. Jaccard index of centrality measures in the climax phase**

| Measure | jacc | P(<=Jacc) | P(>=Jacc) |
| --- | --- | --- | --- |
| degree | 0.38 | 0.77 | 0.35 |
| betweenness centr. | 0.19 | 0.09 | 0.96 |
| closeness centr. | 0.28 | 0.37 | 0.76 |
| eigenvec. centr. | 0.38 | 0.77 | 0.35 |
| hub taxa | 0.00 * | 0.03 * | 1.00 |

**Table S5.** **Network comparison in the attack phase**

|  | Sputum | Throat_s | Abs_diff. | P_value |
| --- | --- | --- | --- | --- |
| **Global network measures** | | | | |
| Relative LCC size | 0.98 | 1.00 | 0.01 | 0.68 |
| Clustering coefficient | 0.55 | 0.60 | 0.05 | 0.32 |
| Modularity | 0.11 | 0.17 | 0.05 | 0.33 |
| Positive edge percentage | 49.08 | 54.13 | 5.05 | 0.04 * |
| Edge density | 0.30 | 0.26 | 0.03 | 0.45 |
| Natural connectivity | 0.07 | 0.08 | 0.00 | 0.73 |
| Vertex connectivity | 3.00 | 2.00 | 1.00 | 0.84 |
| Edge connectivity | 3.00 | 2.00 | 1.00 | 0.84 |
| Average dissimilarity* | 0.89 | 0.90 | 0.00 | 0.82 |
| Average path length | 1.02 | 1.13 | 0.10 | 0.08 . |
| **Degree (weighted)** | | | | |
| *Schaalia* JVLH_s | 0.00 | 12.490 | 12.49 | 0.41 |
| *Haemophilus haemolyticus* | 9.67 | 2.103 | 7.57 | 0.98 |
| *Gemella haemolysan* | 8.57 | 2.680 | 5.89 | 0.98 |
| *Campylobacter gracilis* | 9.36 | 4.010 | 5.35 | 0.98 |
| *Haemophilus parainfluenzae* | 7.04 | 12.060 | 5.01 | 0.98 |
| *Eikenella corrodens* | 5.24 | 0.990 | 4.25 | 0.98 |
| **Eigenvector centrality (normalized)** | | | | |
| *Gemella haemolysans* | 0.78 | 0.05 | 0.72 | 0.65 |
| *Schaalia* JVLH_s | 0.00 | 0.61 | 0.61 | 0.65 |
| *Campylobacter gracilis* | 0.86 | 0.28 | 0.58 | 0.65 |
| *Haemophilus haemolyticus* | 0.64 | 0.10 | 0.54 | 0.65 |
| *Rothia* KV795237_s | 0.64 | 0.10 | 0.53 | 0.65 |
| *Streptococcus_sp* | 0.64 | 0.10 | 0.53 | 0.65 |

**Table S6. Jaccard index of centrality measures in the attack phase**

| Measure | jacc | P(<=Jacc) | P(>=Jacc) |
| --- | --- | --- | --- |
| degree | 0.33 | 0.58 | 0.57 |
| betweenness centr. | 0.58 | 0.03 * | 0.98 |
| closeness centr. | 0.38 | 0.77 | 0.35 |
| eigenvec. centr. | 0.38 | 0.77 | 0.35 |
| hub taxa | 0.00 | 0.03 * | 0.10 |
